# Attitudes towards deprescribing in patients with multimorbidity and polypharmacy in primary care

**DOI:** 10.1101/2024.12.19.24319303

**Authors:** Eduard Shantsila, Alan Woodall, Frances S Mair, Aseel S Abuzour, Danushka Bollegala, Harriet Cant, Andrew Clegg, Mark Gabbay, Alan Griffiths, Layik Hama, James Harmsworth-King, Benedict Jones, Gary Leeming, Emma Lo, Simon Maskell, Maurice O’Connell, Olusegun Popoola, Samuel Relton, Asra Aslam, Roy A Ruddle, Pieta Schofield, Matthew Sperrin, Tjeerd Van Staa, Samantha Wilson, Iain Buchan, Lauren E Walker

**Author notes:** joint first authors (Eduard Shantsila, Alan Woodall). **Corresponding author:** Lauren Walker.

## Abstract

**Background:** Population ageing has led to an increase in multimorbidity and polypharmacy. Some medications may need to be stopped, but patient attitudes towards deprescribing are poorly understood. This study explores attitudes towards (de)prescribing in patients with multimorbidity in the UK primary care.

**Methods:** Patients with multimorbidity were invited to complete the Revised Patients’ Attitudes Towards Deprescribing (rPATD) Questionnaire using an Evergreen Life’s Personal Health Record App (Manchester, UK). The responses were linked to electronic health records. Anonymised data were analysed in a trusted research environment (University of Liverpool) for group comparisons and using multivariable logistic regression to identify factors associated with satisfaction with current medications.

**Results:** A total 1,019 patients participated in the study (n=365 aged <65, 30% males; n=654 ≥65, 57% males). Most patients were satisfied with their current medications (74% aged <65, 70% aged ≥65) but were willing to stop one or more of their regular medicines if their doctor said it was possible (82%, 68% accordingly). Polypharmacy, use of antihypertensive drugs, and antidepressants were associated with patient-reported burden in taking medicines. Frailty did not influence patient deprescribing attitudes. Patients who were satisfied with current medications had fewer medications. Independent predictors of satisfaction with current medications were higher total involvement and appropriateness scores, and lower total burden score.

**Conclusions:** Most patients with multimorbidity would consider stopping some of their medications, even when they are generally satisfied with the treatments they received. Frailty status does not imply willingness to stop medications. Clinicians should discuss medication deprescribing for shared decision.

## Introduction

Ageing populations, changes in lifestyle risk factors, and medical advances are associated with an increase in multimorbidity, wherein people live with two or more long-term conditions, often leading to polypharmacy.(1) The prevalence of polypharmacy in the UK is increasing. In England, for example, the number of medications dispensed in primary care increased by 53.8% between 2001 and 2011.(2) While many medications improve survival and quality of life, polypharmacy (using ≥5 regular medications) is associated with increased risk of medicine-related harm, including hospital admission, poorer quality of life, and mortality.(3) Deprescribing is defined as supervised discontinuation or dose reduction of one or more medications which are considered clinically inappropriate.(4) Deprescribing may decrease medication burden and risks of medication-related harm such as hospital admission which accrue with polypharmacy(5), and improve health outcomes in selected patients, particularly the frail and elderly.(6) In 2021, the UK government commissioned an independent review in England to develop a plan to reduce inappropriate prescribing. This highlighted a significant increase in NHS prescribing, with an increase in medication costs and the negative impact on patients and society. An individual taking ten or more regular medicines is three times more likely to be admitted to hospital because of an adverse drug reaction (ADR).(7) Primary care is a major source of prescribing, responsible for more than one billion prescription items dispensed annually in the UK, with an estimated 10% being inappropriately prescribed.(8) A number of factors decrease the confidence of clinicians to deprescribe: the information for coordinating care is hard to assemble and understand, particularly in time-constrained consultations; there is often lack of appropriate guidance for complex patients with multimorbidity, with the majority of guidelines largely based on single-condition research evidence; and clinicians voice concerns regarding potential patient conflicts and attitudes toward deprescribing.(9) Understanding patient views toward deprescribing is thus essential for better education strategies, efficient communication from clinicians and compliance with medication.

There is limited information on public views on deprescribing in the UK. The Revised Patients’ Attitudes Towards Deprescribing (rPATD) Questionnaire: Versions for Older Adults and Caregivers tool has been developed and validated to understand deprescribing attitudes in older adults and has been used in different countries.(10) Of particular interest would be information on how attitudes to deprescribing are affected by those at the greatest risk of medicines related harm in primary care: these include patients who have complex multimorbidity (defined as ≥4 long-term health conditions plus ≥10 medicines); those aged ≥65 years with frailty, and those who have co-morbid physical and mental illness, especially those taking antidepressants and antipsychotics.(11, 12) This study explores, for the first time, the attitudes towards prescribing in patients with multimorbidity and polypharmacy associated with high-risk of medicines harm, in the UK primary care setting.

## Methods

Eligible registered users of the Evergreen Life’s Personal Health Record (PHR) App (Manchester, UK) were invited to participate in a survey on their attitudes to deprescribing using the rPATD questionnaire (Supplemental Table 1).(10) The Evergreen Life PHR App links health information, including access to NHS-assured GP services, personal health and wellbeing records. App users can consent for personal data, including through responses to wellness surveys, to be used in specific research studies. Following consent and completion of a survey, Evergreen Life anonymises and aggregates data for research purposes. Our study received ethical approval from North-East – Newcastle & North Tyneside Research Ethics Committee (REC reference 22/NE/0088). Offers to participate in the survey were sent through the app three occasions, a minimum of 72 hours apart. It was acceptable for participants to not respond to some questions.

Eligible participants were existing users of Evergreen Life PHR App Adults aged ≥18 years with multimorbidity defined as ≥2 long-term conditions who consented for participation in the survey. Multimorbidity was defined according to the Academy of Medical Sciences definition as co-existence of ≥2 chronic conditions, each one of which is either a physical non-communicable disease of long duration (e.g., cardiovascular disease, diabetes mellitus, cancer), a mental health condition of long duration (e.g., a mood disorder, schizophrenia, dementia), or an infectious disease of long duration (e.g., HIV or hepatitis C). A disease code list for 186 chronic health conditions was developed by a group of general physicians. Frailty was defined using electronic Frailty index (eFI). Polypharmacy was defined as use of ≥5 medications issued ≥4 times within a 12-month period.

Participants were asked to score their rPATD answers using a score range from strongly disagree (1) to strongly agree (5). Average scores were calculated in four domains of five questions each, based on the person’s own perception: total involvement score, with higher scores indicating more patient involvement with their medicines/deprescribing; total burden score, with higher scores indicating them seeing their medications more burdensome; total appropriateness score, with higher scores indicating the patients viewed their medications more appropriate; and total concerns about stopping score, with higher scores indicating more potential concerns the patients had about stopping one or more of their medications. Additionally, two questions were global questions not included in any scores: ‘Overall, I am satisfied with my current medicines’ and ‘If my doctor said it was possible I would be willing to stop one or more of my regular medicines.’ The rPATD questionnaire responses were collated and cross-referenced against the pseudo-anonymised patient demographic and clinical characteristics. Following this data were fully anonymised prior to analyses conduced within a Trustworthy Research Environment at the University of Liverpool. We were particularly interested in the relationship between antidepressant and antihypertensive use and attitudes towards deprescribing due to the high prevalence of use of these medications in the population.

### Statistical analysis

Continuous data are summarised as means and standard deviations (SD) with independent groups compared using a two-sample Student t-test. Categorical data are summarised as the number of survey respondents and the percentage giving specific responses, and associations between categories were compared using Fisher test. Logistic regression was used to explore factors associated with satisfaction with current medications. P-values <0.05 were considered statistically significant. Analyses were undertaken using R software (version 4.3, using R Markdown 2024, tidyverse, epitools, finalfit packages, reproducible table tables produced using gt summary package).

## Results

A total of 11,765 eligible patients were invited to participate in the survey, of whom 1,019 participated (response rate of 8.7%). They included 365 patients aged <65 years (mean (SD) age 51 (10) years, 30% males, 91% of White ethnic origin) and 654 patients aged ≥65 years (age 74 (6) years, 57% males, 96% of White ethnic origin). The age, sex and ethnicity profiles of those invited vs participating in the survey were similar. Demographic and clinical characteristics of study participants are summarized in Table 1.

**Table 1.** Demographic and clinical characteristics of study participants.

| Characteristic | N | <65 years<br>n = 365 | ≥65 years<br>n = 654 | p |
| --- | --- | --- | --- | --- |
| Age, years, Mean (SD) | 1,019 | 51 (10) | 74 (6) | <0.001 |
| Male sex, n (%) | 1,017 | 109 (30%) | 372 (57%) | <0.001 |
| White ethnicity, n (%) | 584 | 213 (91%) | 335 (96%) | 0.023 |
| Body mass index, kg/m<sup>2</sup>, Mean (SD) | 989 | 32.2 (7.6) | 28.1 (5.3) | <0.001 |
| History of smoking, n (%) | 614 | 138 (58%) | 228 (61%) | 0.55 |
| Alcohol use, unis/month, Mean (SD) | 384 | 16 (31) | 21 (34) | 0.015 |
| Frailty, n (%) | 575 | - | 248 (43%) | - |
| Bone fracture, n (%) | 1,019 | 67 (18%) | 86 (13%) | 0.028 |
| Hypertension, n (%) | 1,019 | 100 (27%) | 307 (47%) | <0.001 |
| Myocardial infarction, n (%) | 1,019 | 6 (1.6%) | 20 (3.1%) | 0.22 |
| Heart failure, n (%) | 1,019 | 1 (0.3%) | 21 (3.2%) | 0.001 |
| COPD, n (%) | 1,019 | 7 (1.9%) | 41 (6.3%) | 0.001 |
| Diabetes type 1, n (%) | 1,019 | 10 (2.7%) | 6 (0.9%) | 0.034 |
| Diabetes type 2, n (%) | 1,019 | 59 (16%) | 102 (16%) | 0.86 |
| Chronic kidney disease, n (%) | 1,019 | 10 (2.7%) | 81 (12%) | <0.001 |
| Dementia, n (%) | 1,019 | 1 (0.3%) | 2 (0.3%) | >0.99 |
| Count of currently prescribed medications, Mean (SD) | 1,019 | 7.5 (4.4) | 6.9 (3.9) | 0.043 |
| Polypharmacy, n (%) | 1,019 | 262 (72%) | 456 (70%) | 0.52 |
| Antihypertensive medications, n (%) | 1,019 | 210 (58%) | 499 (76%) | <0.001 |
| Antidepressants, n (%) | 1,019 | 266 (73%) | 211 (32%) | <0.001 |
| Antipsychotic medications, n (%) | 1,019 | 28 (7.7%) | 5 (0.8%) | <0.001 |
| High anticholinergic burden, n (%) | 873 | 264 (77%) | 342 (65%) | <0.001 |
| Anticholinergic burden, Mean (SD) | 873 | 6.9 (5.7) | 4.4 (3.5) | <0.001 |
| Overall, I am satisfied with my current medicines, n (%) | 1,001 |  |  | 0.006 |
| Strongly agree |  | 69 (19%) | 130 (20%) |  |
| Agree |  | 184 (51%) | 345 (54%) |  |
| Unsure |  | 76 (21%) | 142 (22%) |  |
| Disagree |  | 27 (7.5%) | 16 (2.5%) |  |
| Strongly disagree |  | 6 (1.7%) | 6 (0.9%) |  |
| If my doctor said it was possible I would be willing to stop one or more of my regular | 865 |  |  | 0.001 |
| medicines, n (%) |  |  |  |  |
| Strongly agree |  | 90 (31%) | 200 (35%) |  |
| Agree |  | 107 (37%) | 269 (47%) |  |
| Unsure |  | 61 (21%) | 80 (14%) |  |
| Disagree |  | 18 (6.3%) | 19 (3.3%) |  |
| Strongly disagree |  | 11 (3.8%) | 10 (1.7%) |  |
| Total involvement score,<br>Mean (SD) | 971 | 4.40 (0.51) | 4.31 (0.49) | 0.011 |
| Total burden score, Mean<br>(SD) | 869 | 2.66 (0.84) | 2.41 (0.78) | <0.001 |
| Total appropriateness score,<br>Mean (SD) | 842 | 3.46 (0.83) | 3.35 (0.80) | 0.065 |
| Total concerns about stop-<br>ping score, Mean (SD) | 812 | 2.95 (0.84) | 2.58 (0.69) | <0.001 |

Overall, patients in the older group were more satisfied with their current medications (74% vs 70%, p=0.006) and would be willing to stop one or more of their regular medicines if their doctor said it was possible (82% vs 68%, p=0.001) (Table 1). The older groups showed lower total involvement scores, total burden scores and total concerns about stopping scores (p<0.05 for all). The mean number of currently prescribed medications was 7.5 (SD 4.4) in those aged <65 and 6.9 (SD 3.9) in those aged ≥65.Most participants were satisfied with their current medications (either satisfied or very satisfied survey choice), 70% aged <65, and 74% aged ≥65. Still, most would be willing to stop one or more of their regular medicines if their doctor said it was possible (68% aged <65, 82% aged ≥65). The willingness to stop medication(s) was highest in those aged ≥65 if receiving antihypertensive drugs. ( see Figure 1 and Table 7).

**Figure 1.**
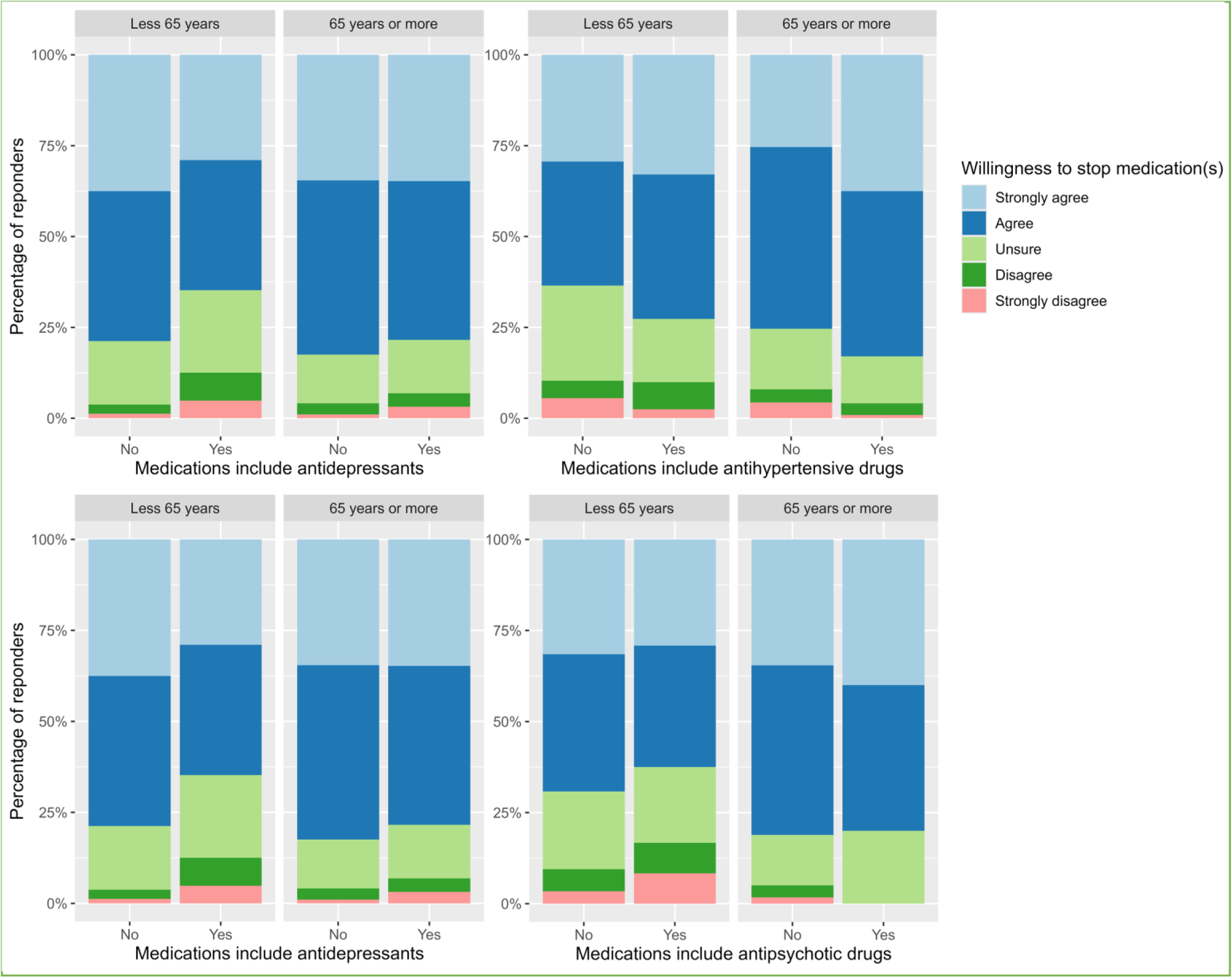
Willingness to stop medications in the study groups.

### Polypharmacy

In patients aged <65, those with polypharmacy (≥5 regular medications, n=262) had a higher mean total burden score of 2.74 (SD 0.86) compared to those without polypharmacy (n=103, mean score of 2.41 (SD 0.72), p=0.001). In patients aged ≥65, those with polypharmacy (n=456) have increased total burden score, total concerns about stopping score and lower total appropriateness score compared to those without polypharmacy (n=198, p<0.001 for all) (Table 2).

**Table 2.** Effect of polypharmacy (5 or more regular medications) on attitudes to deprescribing.

| Characteristic | Polypharmacy: <65 years |  |  |  | Polypharmacy: ≥65 years |  |  |  |
| --- | --- | --- | --- | --- | --- | --- | --- | --- |
|  | N | No n=103 | Yes n=262 | p | N | No n=198 | Yes n=456 | p |
| Age, years, Mean (SD) | 365 | 49 (11) | 51 (10) | 0.020 | 654 | 73 (5) | 74 (6) | 0.004 |
| Male sex, n (%), n (%) | 364 | 30 (29%) | 79 (30%) | 0.90 | 653 | 118 (60%) | 254 (56%) | 0.39 |
| White ethnicity, n (%) | 234 | 52 (91%) | 161 (91%) | >0.99 | 350 | 106 (98%) | 229 (95%) | 0.16 |
| Body mass index, kg/m<sup>2</sup>, Mean (SD) | 348 | 30 (6) | 33 (8) | <0.001 | 641 | 27.2 (4.9) | 28.5 (5.5) | 0.002 |
| History of smoking, n (%) | 238 | 32 (52%) | 106 (60%) | 0.29 | 376 | 72 (65%) | 156 (59%) | 0.30 |
| Alcohol use, unis/month, Mean (SD) | 165 | 24 (39) | 13 (28) | 0.13 | 219 | 25 (37) | 19 (32) | 0.28 |
| Frailty, n (%) | 0 | 0 (NA%) | 0 (NA%) |  | 575 | 69 (36%) | 179 (46%) | 0.025 |
| Bone fracture, n (%) | 365 | 16 (16%) | 51 (19%) | 0.45 | 654 | 27 (14%) | 59 (13%) | 0.80 |
| Hypertension, n (%) | 365 | 17 (17%) | 83 (32%) | 0.004 | 654 | 76 (38%) | 231 (51%) | 0.005 |
| Myocardial infarction, n (%) | 365 | 0 (0%) | 6 (2.3%) | 0.19 | 654 | 0 (0%) | 20 (4.4%) | <0.001 |
| Heart failure, n (%) | 365 | 0 (0%) | 1 (0.4%) | >0.99 | 654 | 2 (1.0%) | 19 (4.2%) | 0.050 |
| COPD, n (%) | 365 | 0 (0%) | 7 (2.7%) | 0.20 | 654 | 6 (3.0%) | 35 (7.7%) | 0.023 |
| Diabetes type 1, n (%) | 365 | 0 (0%) | 10 (3.8%) | 0.068 | 654 | 0 (0%) | 6 (1.3%) | 0.19 |
| Diabetes type 2, n (%) | 365 | 8 (7.8%) | 51 (19%) | 0.007 | 654 | 16 (8.1%) | 86 (19%) | <0.001 |
| Chronic kidney disease, n (%) | 365 | 3 (2.9%) | 7 (2.7%) | >0.99 | 654 | 17 (8.6%) | 64 (14%) | 0.053 |
| Dementia, n (%) | 365 | 1 (1.0%) | 0 (0%) | 0.28 | 654 | 0 (0%) | 2 (0.4%) | >0.99 |
| Count of currently prescribed medications, Mean (SD) | 365 | 3.1 (0.8) | 9.2 (4.0) | <0.001 | 654 | 3.1 (0.8) | 8.6 (3.5) | <0.001 |
| Antihypertensive medications, n (%) | 365 | 46 (45%) | 164 (63%) | 0.002 | 654 | 115 (58%) | 384 (84%) | <0.001 |
| Antidepressants, n (%) | 365 | 69 (67%) | 197 (75%) | 0.12 | 654 | 37 (19%) | 174 (38%) | <0.001 |
| Antipsychotic medications, n (%) | 365 | 4 (3.9%) | 24 (9.2%) | 0.12 | 654 | 1 (0.5%) | 4 (0.9%) | >0.99 |
| High anticholinergic burden, n (%) | 343 | 47 (55%) | 217 (84%) | <0.001 | 530 | 52 (45%) | 290 (70%) | <0.001 |
| Anticholinergic burden, Mean (SD) | 343 | 3.8 (2.7) | 7.9 (6.0) | <0.001 | 530 | 2.9 (2.5) | 4.8 (3.7) | <0.001 |
| Overall, I am satisfied with my current medicines, n (%) | 362 |  |  | 0.20 | 639 |  |  | 0.007 |
| Strongly agree |  | 21 (21%) | 48 (18%) |  |  | 45 (23%) | 85 (19%) |  |
| Agree |  | 59 (58%) | 125 (48%) |  |  | 117 (60%) | 228 (51%) |  |
| Unsure |  | 16 (16%) | 60 (23%) |  |  | 30 (15%) | 112 (25%) |  |
| Disagree |  | 6 (5.9%) | 21 (8.1%) |  |  | 2 (1.0%) | 14 (3.1%) |  |
| Strongly disagree |  | 0 (0%) | 6 (2.3%) |  |  | 0 (0%) | 6 (1.3%) |  |
| If my doctor said it was possible I would be willing to stop one or more of my regular medicines, n (%) | 287 |  |  | 0.93 | 578 |  |  | 0.92 |
| Strongly agree |  | 23 (32%) | 67 (31%) |  |  | 55 (33%) | 145 (35%) |  |
| Agree |  | 27 (37%) | 80 (37%) |  |  | 81 (48%) | 188 (46%) |  |
| Unsure |  | 15 (21%) | 46 (21%) |  |  | 25 (15%) | 55 (13%) |  |
| Disagree |  | 4 (5.5%) | 14 (6.5%) |  |  | 6 (3.6%) | 13 (3.2%) |  |
| Strongly disagree |  | 4 (5.5%) | 7 (3.3%) |  |  | 2 (1.2%) | 8 (2.0%) |  |
| Total involvement score, Mean (SD) | 344 | 4.39 (0.48) | 4.40 (0.52) | 0.90 | 627 | 4.32 (0.54) | 4.30 (0.47) | 0.78 |
| Total burden score, Mean (SD) | 287 | 2.41 (0.72) | 2.74 (0.86) | 0.001 | 582 | 2.04 (0.67) | 2.57 (0.77) | <0.001 |
| Total appropriateness score, Mean (SD) | 277 | 3.59 (0.83) | 3.42 (0.83) | 0.13 | 565 | 3.66 (0.71) | 3.22 (0.80) | <0.001 |
| Total concerns about stopping score, Mean (SD) | 268 | 2.89 (0.81) | 2.98 (0.85) | 0.46 | 544 | 2.39 (0.68) | 2.66 (0.68) | <0.001 |

### Antihypertensive medications

Antihypertensive drugs were used by 210 (56%) patients aged <65, and 499 (76%) of those aged ≥65 (Table 3). Patients aged <65 years who used antihypertensive drugs had a higher total burden score vs those without these drugs (p=0.036). Among patients aged ≥65, those receiving antihypertensive drugs were more willing to stop one or more of their regular medicines (83% vs 75% in those not receiving antihypertensive medication, p=0.011), had a higher total burden score and lower total appropriateness score (p<0.001 for both).

**Table 3.** Effect of antihypertensives drug use on attitudes to deprescribing.

| Characteristic | Antihypertensive drugs: <65 years |  |  |  | Antihypertensive drugs: ≥65 years |  |  |  |
| --- | --- | --- | --- | --- | --- | --- | --- | --- |
|  | N | No<br>n = 155 | Yes<br>n = 210 | p | N | No<br>n = 155 | Yes<br>n = 499 | p |
| Age, years, Mean (SD) | 365 | 49 (10) | 52 (10) | 0.025 | 654 | 73 (6) | 74 (6) | 0.11 |
| Male sex, n (%), n (%) | 364 | 40 (26%) | 69 (33%) | 0.16 | 653 | 69 (45%) | 303 (61%) | <0.001 |
| White ethnicity, n (%) | 234 | 90 (91%) | 123 (91%) | >0.99 | 350 | 77 (99%) | 258 (95%) | 0.21 |
| Body mass index, kg/m<sup>2</sup>, Mean (SD) | 348 | 31 (7) | 33 (8) | 0.036 | 641 | 26.4 (4.6) | 28.6 (5.4) | <0.001 |
| History of smoking, n (%) | 238 | 57 (59%) | 81 (57%) | 0.89 | 376 | 54 (63%) | 174 (60%) | 0.71 |
| Alcohol use, unis/month, Mean (SD) | 165 | 11 (25) | 18 (34) | 0.12 | 219 | 20 (33) | 21 (34) | 0.86 |
| Frailty, n (%) | 0 | 0 (NA%) | 0 (NA%) |  | 575 | 51 (38%) | 197 (45%) | 0.14 |
| Bone fracture, n (%) | 365 | 28 (18%) | 39 (19%) | >0.99 | 654 | 27 (17%) | 59 (12%) | 0.078 |
| Hypertension, n (%) | 365 | 3 (1.9%) | 97 (46%) | <0.001 | 654 | 8 (5.2%) | 299 (60%) | <0.001 |
| Myocardial infarction, n (%) | 365 | 0 (0%) | 6 (2.9%) | 0.041 | 654 | 1 (0.6%) | 19 (3.8%) | 0.058 |
| Heart failure, n (%) | 365 | 0 (0%) | 1 (0.5%) | >0.99 | 654 | 1 (0.6%) | 20 (4.0%) | 0.037 |
| COPD, n (%) | 365 | 2 (1.3%) | 5 (2.4%) | 0.70 | 654 | 14 (9.0%) | 27 (5.4%) | 0.13 |
| Diabetes type 1, n (%) | 365 | 3 (1.9%) | 7 (3.3%) | 0.53 | 654 | 2 (1.3%) | 4 (0.8%) | 0.63 |
| Diabetes type 2, n (%) | 365 | 18 (12%) | 41 (20%) | 0.045 | 654 | 12 (7.7%) | 90 (18%) | 0.001 |
| Chronic kidney disease, n (%) | 365 | 2 (1.3%) | 8 (3.8%) | 0.20 | 654 | 10 (6.5%) | 71 (14%) | 0.011 |
| Dementia, n (%) | 365 | 1 (0.6%) | 0 (0%) | 0.42 | 654 | 0 (0%) | 2 (0.4%) | >0.99 |
| Count of currently prescribed medications, Mean (SD) | 365 | 6.4 (3.9) | 8.3 (4.6) | <0.001 | 654 | 5.2 (3.3) | 7.4 (3.9) | <0.001 |
| Polypharmacy, n (%) | 365 | 98 (63%) | 164 (78%) | 0.002 | 654 | 72 (46%) | 384 (77%) | <0.001 |
| antidepressants, n (%) | 365 | 116 (75%) | 150 (71%) | 0.48 | 654 | 56 (36%) | 155 (31%) | 0.24 |
| Antipsychotic medications, n (%) | 365 | 17 (11%) | 11 (5.2%) | 0.048 | 654 | 3 (1.9%) | 2 (0.4%) | 0.089 |
| High anticholinergic burden, n (%) | 343 | 104 (71%) | 160 (81%) | 0.038 | 530 | 73 (60%) | 269 (66%) | 0.24 |
| Anticholinergic burden, Mean (SD) | 343 | 6.0 (5.2) | 7.5 (6.0) | 0.019 | 530 | 4.3 (3.8) | 4.4 (3.5) | 0.91 |
| Overall, I am satisfied with my current medicines, n (%) | 362 |  |  | 0.055 | 639 |  |  | 0.22 |
| Strongly agree |  | 39 (25%) | 30 (14%) |  |  | 34 (23%) | 96 (20%) |  |
| Agree |  | 68 (44%) | 116 (56%) |  |  | 85 (56%) | 260 (53%) |  |

|  | Antihypertensive drugs: <65 years |  |  |  | Antihypertensive drugs: ≥65 years |  |  |  |
| --- | --- | --- | --- | --- | --- | --- | --- | --- |
| Characteristic | N | No<br>n = 155 | Yes<br>n = 210 | p | N | No<br>n = 155 | Yes<br>n = 499 | p |
| Unsure |  | 31 (20%) | 45 (22%) |  |  | 25 (17%) | 117 (24%) |  |
| Disagree |  | 14 (9.1%) | 13 (6.3%) |  |  | 6 (4.0%) | 10 (2.0%) |  |
| Strongly disagree |  | 2 (1.3%) | 4 (1.9%) |  |  | 1 (0.7%) | 5 (1.0%) |  |
| If my doctor said it was possible I would be willing to stop one or more of my regular medicines, n (%) | 287 |  |  | 0.20 | 578 |  |  | 0.011 |
| Strongly agree |  | 37 (29%) | 53 (33%) |  |  | 35 (25%) | 165 (38%) |  |
| Agree |  | 43 (34%) | 64 (40%) |  |  | 69 (50%) | 200 (45%) |  |
| Unsure |  | 33 (26%) | 28 (17%) |  |  | 23 (17%) | 57 (13%) |  |
| Disagree |  | 6 (4.8%) | 12 (7.5%) |  |  | 5 (3.6%) | 14 (3.2%) |  |
| Strongly disagree |  | 7 (5.6%) | 4 (2.5%) |  |  | 6 (4.3%) | 4 (0.9%) |  |
| Total involvement score, Mean (SD) | 344 | 4.45 (0.51) | 4.35 (0.51) | 0.071 | 627 | 4.36 (0.45) | 4.29 (0.51) | 0.15 |
| Total burden score, Mean (SD) | 287 | 2.54 (0.82) | 2.75 (0.84) | 0.036 | 582 | 2.17 (0.73) | 2.49 (0.78) | <0.001 |
| Total appropriateness score, Mean (SD) | 277 | 3.52 (0.85) | 3.42 (0.82) | 0.35 | 565 | 3.58 (0.81) | 3.28 (0.78) | <0.001 |
| Total concerns about stopping score, Mean (SD) | 268 | 2.98 (0.82) | 2.94 (0.85) | 0.71 | 544 | 2.59 (0.74) | 2.58 (0.68) | 0.93 |

### Antidepressants

Antidepressant drugs were used by 266 (73%) patients aged <65, and 211 (32%) of those aged ≥65 years (Table 4). Patients aged <65 years who used antidepressant drugs had a lower total appropriateness score (p=0.041) and a higher total concern about stopping score (<0.001). Among patients aged ≥65, fewer of those receiving antidepressants were satisfied with their current medications (68% vs 78% of those without antidepressants, p<0.001). Patients aged ≥65 using antidepressants had a higher total burden score (p=0.002), a lower total appropriateness score (p=0.024), and a higher total concern about stopping score (p<0.001). The effects of antipsychotic medications were not analysed separately due to the small number of participants receiving these drugs (n=28, aged <65, n=5, aged ≥65).

**Table 4.** Effect of antidepressant use on attitudes to deprescribing.

| Characteristic | Antidepressants: <65 years |  |  |  | Antidepressants: ≥65 years |  |  |  |
| --- | --- | --- | --- | --- | --- | --- | --- | --- |
|  | N | No<br>n = 99 | Yes<br>n = 266 | p | N | No<br>n = 443 | Yes<br>n = 211 | p |
| Age, years, Mean (SD) | 365 | 53 (9) | 50 (10) | 0.001 | 654 | 74 (6) | 73 (6) | 0.006 |
| Male sex, n (%), n (%) | 364 | 44 (45%) | 65 (24%) | <0.001 | 653 | 283 (64%) | 89 (42%) | <0.001 |
| White ethnicity, n (%) | 234 | 42 (86%) | 171 (92%) | 0.16 | 350 | 223 (95%) | 112 (97%) | 0.78 |
| Body mass index, kg/m<sup>2</sup>, Mean (SD) | 348 | 30 (7) | 33 (8) | <0.001 | 641 | 27.6 (4.8) | 29.1 (6.1) | 0.003 |
| History of smoking, n (%) | 238 | 28 (48%) | 110 (61%) | 0.094 | 376 | 145 (58%) | 83 (65%) | 0.22 |
| Alcohol use, unis/month, Mean (SD) | 165 | 17 (27) | 15 (33) | 0.79 | 219 | 20 (33) | 21 (36) | 0.85 |
| Frailty, n (%) | 0 | 0 (NA%) | 0 (NA%) |  | 575 | 177 (44%) | 71 (42%) | 0.71 |
| Bone fracture, n (%) | 365 | 18 (18%) | 49 (18%) | >0.99 | 654 | 53 (12%) | 33 (16%) | 0.22 |
| Hypertension, n (%) | 365 | 32 (32%) | 68 (26%) | 0.23 | 654 | 205 (46%) | 102 (48%) | 0.68 |
| Myocardial infarction, n (%) | 365 | 4 (4.0%) | 2 (0.8%) | 0.048 | 654 | 12 (2.7%) | 8 (3.8%) | 0.47 |
| Heart failure, n (%) | 365 | 1 (1.0%) | 0 (0%) | 0.27 | 654 | 13 (2.9%) | 8 (3.8%) | 0.64 |
| COPD, n (%) | 365 | 1 (1.0%) | 6 (2.3%) | 0.68 | 654 | 22 (5.0%) | 19 (9.0%) | 0.057 |
| Diabetes type 1, n (%) | 365 | 5 (5.1%) | 5 (1.9%) | 0.14 | 654 | 3 (0.7%) | 3 (1.4%) | 0.39 |
| Diabetes type 2, n (%) | 365 | 23 (23%) | 36 (14%) | 0.037 | 654 | 72 (16%) | 30 (14%) | 0.56 |
| Chronic kidney disease, n (%) | 365 | 2 (2.0%) | 8 (3.0%) | >0.99 | 654 | 58 (13%) | 23 (11%) | 0.45 |
| Dementia, n (%) | 365 | 0 (0%) | 1 (0.4%) | >0.99 | 654 | 1 (0.2%) | 1 (0.5%) | 0.54 |
| Count of currently prescribed medications, Mean (SD) | 365 | 6.8 (4.1) | 7.7 (4.5) | 0.069 | 654 | 6.1 (3.4) | 8.6 (4.4) | <0.001 |
| Polypharmacy, n (%) | 365 | 65 (66%) | 197 (74%) | 0.12 | 654 | 282 (64%) | 174 (82%) | <0.001 |
| Antihypertensive medications, n (%) | 365 | 60 (61%) | 150 (56%) | 0.48 | 654 | 344 (78%) | 155 (73%) | 0.24 |
| Antipsychotic medications, n (%) | 365 | 0 (0%) | 28 (11%) | <0.001 | 654 | 3 (0.7%) | 2 (0.9%) | 0.66 |
| High anticholinergic burden, n (%) | 343 | 43 (55%) | 221 (83%) | <0.001 | 530 | 155 (48%) | 187 (89%) | <0.001 |
| Anticholinergic burden, Mean (SD) | 343 | 3.3 (2.4) | 7.9 (5.9) | <0.001 | 530 | 3.0 (2.4) | 6.4 (4.0) | <0.001 |
| Overall, I am satisfied with my current medicines, n (%) | 362 |  |  | 0.088 | 639 |  |  | <0.001 |
| Strongly agree |  | 26 (27%) | 43 (16%) |  |  | 99 (23%) | 31 (15%) |  |
| Agree |  | 49 (50%) | 135 (51%) |  |  | 237 (55%) | 108 (53%) |  |
| Unsure |  | 15 (15%) | 61 (23%) |  |  | 91 (21%) | 51 (25%) |  |
| Disagree |  | 8 (8.2%) | 19 (7.2%) |  |  | 3 (0.7%) | 13 (6.3%) |  |
| Strongly disagree |  | 0 (0%) | 6 (2.3%) |  |  | 4 (0.9%) | 2 (1.0%) |  |
| If my doctor said it was possible I would be willing to stop one or more of my regular medicines, n (%) | 287 |  |  | 0.15 | 578 |  |  | 0.39 |
| Strongly agree |  | 30 (38%) | 60 (29%) |  |  | 134 (35%) | 66 (35%) |  |
| Agree |  | 33 (41%) | 74 (36%) |  |  | 186 (48%) | 83 (44%) |  |
| Unsure |  | 14 (18%) | 47 (23%) |  |  | 52 (13%) | 28 (15%) |  |
| Disagree |  | 2 (2.5%) | 16 (7.7%) |  |  | 12 (3.1%) | 7 (3.7%) |  |
| Strongly disagree |  | 1 (1.3%) | 10 (4.8%) |  |  | 4 (1.0%) | 6 (3.2%) |  |
| Total involvement score, Mean (SD) | 344 | 4.42 (0.53) | 4.38 (0.50) | 0.52 | 627 | 4.32 (0.51) | 4.30 (0.46) | 0.63 |
| Total burden score, Mean (SD) | 287 | 2.55 (0.77) | 2.70 (0.86) | 0.17 | 582 | 2.34 (0.75) | 2.56 (0.82) | 0.002 |
| Total appropriateness score, Mean (SD) | 277 | 3.62 (0.80) | 3.40 (0.84) | 0.041 | 565 | 3.40 (0.79) | 3.24 (0.80) | 0.024 |
| Total concerns about stopping score, Mean (SD) | 268 | 2.57 (0.78) | 3.10 (0.81) | <0.001 | 544 | 2.51 (0.68) | 2.73 (0.69) | <0.001 |

### Frailty

The analysis included 248 (43%) patients with frailty and 327 without frailty, all aged ≥65 years (Table 5). There was no significant difference in attitudes to deprescribing between patients with and without frailty (p>0.05 for all metrics).

**Table 5.**
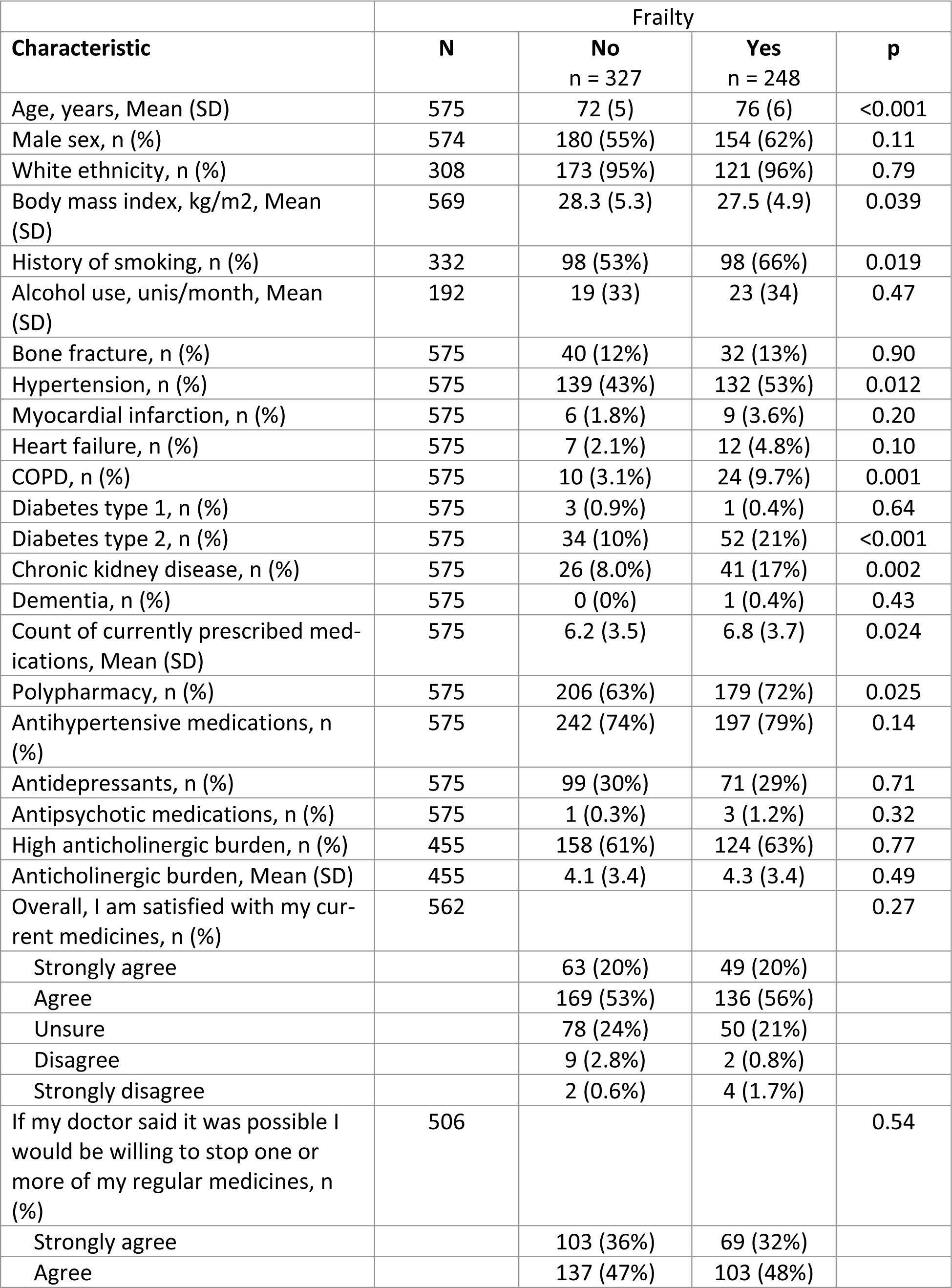
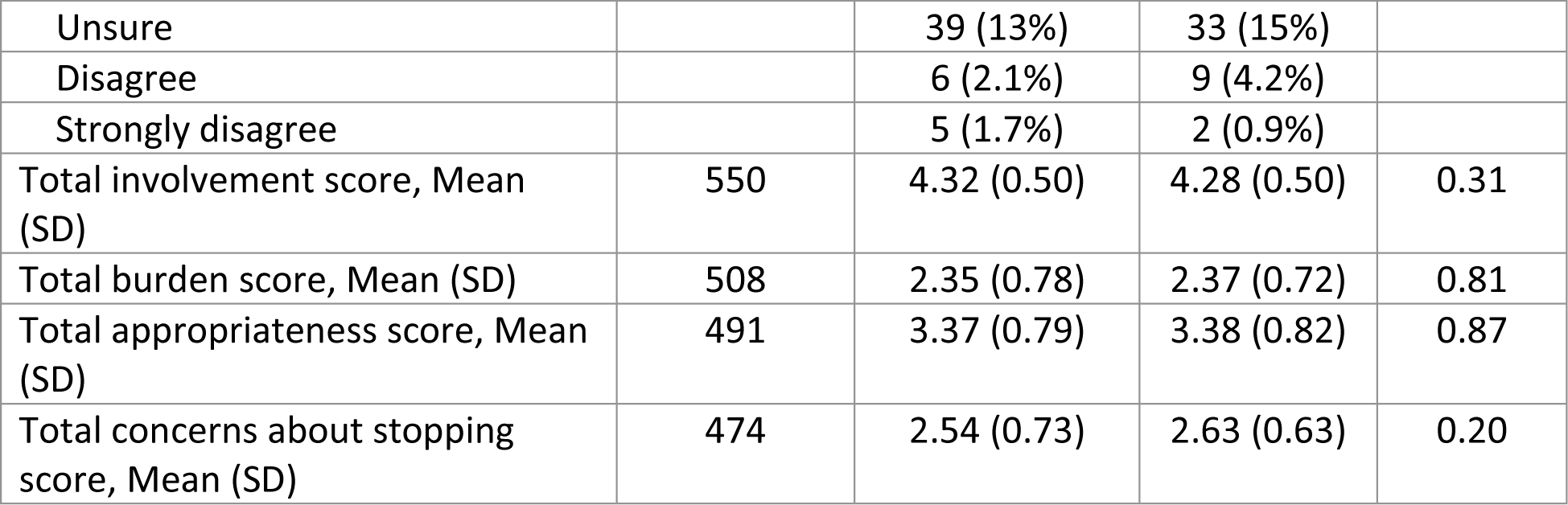
Effect of frailty on attitudes to deprescribing.

### Characteristics associated with satisfaction with current medications

Among total responders 476 patients (70%) were satisfied with current medications (satisfied or very satisfied choice) (Table 6). Patients who were satisfied with current medications had fewer prescribed medications (mean 7.4 (SD 4.0) vs 8.3 (4.1), p=0.007), were less likely to have polypharmacy (75% vs 85%, p=0.004) and less likely to use antidepressants (50% vs 60%, p<0.024). Patients satisfied with current medications had a higher total involvement score, a higher total appropriateness score, and lower total concerns about stopping score (p<0.001 for all). Independent predictors of satisfaction with current medications were a higher total involvement score (p=0.002), a lower total burden score (p=0.028), and a higher total appropriateness score (p<0.001) (Table 7).

**Table 6.**
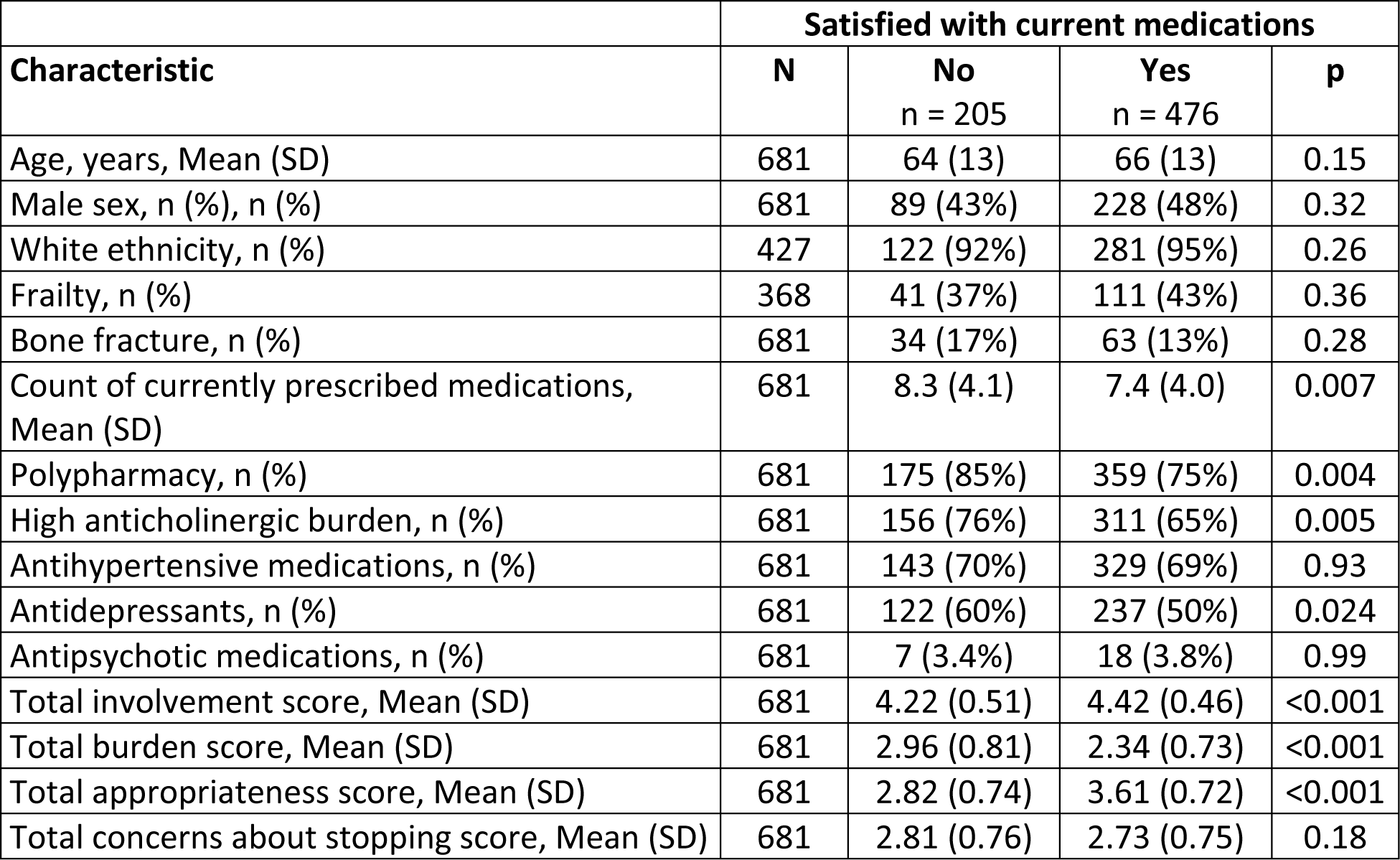
Characteristics of being satisfied/very satisfied with current medications vs lower satisfaction score.

| Characteristic | Satisfied with current medications |  |  |  |
| --- | --- | --- | --- | --- |
|  | N | No<br>n = 205 | Yes<br>n = 476 | p |
| Age, years, Mean (SD) | 681 | 64 (13) | 66 (13) | 0.15 |
| Male sex, n (%), n (%) | 681 | 89 (43%) | 228 (48%) | 0.32 |
| White ethnicity, n (%) | 427 | 122 (92%) | 281 (95%) | 0.26 |
| Frailty, n (%) | 368 | 41 (37%) | 111 (43%) | 0.36 |
| Bone fracture, n (%) | 681 | 34 (17%) | 63 (13%) | 0.28 |
| Count of currently prescribed medications, Mean (SD) | 681 | 8.3 (4.1) | 7.4 (4.0) | 0.007 |
| Polypharmacy, n (%) | 681 | 175 (85%) | 359 (75%) | 0.004 |
| High anticholinergic burden, n (%) | 681 | 156 (76%) | 311 (65%) | 0.005 |
| Antihypertensive medications, n (%) | 681 | 143 (70%) | 329 (69%) | 0.93 |
| Antidepressants, n (%) | 681 | 122 (60%) | 237 (50%) | 0.024 |
| Antipsychotic medications, n (%) | 681 | 7 (3.4%) | 18 (3.8%) | 0.99 |
| Total involvement score, Mean (SD) | 681 | 4.22 (0.51) | 4.42 (0.46) | <0.001 |
| Total burden score, Mean (SD) | 681 | 2.96 (0.81) | 2.34 (0.73) | <0.001 |
| Total appropriateness score, Mean (SD) | 681 | 2.82 (0.74) | 3.61 (0.72) | <0.001 |
| Total concerns about stopping score, Mean (SD) | 681 | 2.81 (0.76) | 2.73 (0.75) | 0.18 |

**Table 7.** Predictors for being satisfied/very satisfied with current medications.

| | Being satisfied/very satisfied with current medications* | | If my doctor said it was possible I would be willing to stop one or more of my regular medicines* | | Total involvement score $\geq 4$ (range 1-5) | | Total appropriateness score $\geq 4$ (range 1-5) | |
| --- | --- | --- | --- | --- | --- | --- | --- | --- |
|  | n=681 |  | n=684 |  | n=690 |  | n=690 |  |
| Characteristic | Odds Ratio (95% CI) | p-value | Odds Ratio (95% CI) | p-value | Odds Ratio (95% CI) | p-value | Odds Ratio (95% CI) | p-value |
| Total involvement score | 1.93 (1.27-2.95) | 0.002 | 1.71 (1.09-2.70) | 0.020 | 1.12 (0.54-2.47) | 0.77 | 1.78 (1.14-2.80) | 0.012 |
| Total burden score | 0.69 (0.50-0.96) | 0.028 | 1.88 (1.29-2.79) | 0.001 | - | - | 0.14 (0.09-0.20) | <0.001 |
| Total appropriateness score | 3.74 (2.67-5.32) | <0.001 | 0.36 (0.25-0.51) | <0.001 | 0.13 (0.07-0.22) | <0.001 | - | - |
| Total concerns about stopping score | 0.95 (0.70-1.28) | 0.73 | 0.34 (0.25-0.47) | <0.001 | 2.09 (1.22-3.66) | 0.009 | 1.29 (0.98-1.71) | 0.072 |
| Age 65 years or more | 1.11 (0.70-1.75) | 0.66 | 1.16 (0.72-1.87) | 0.53 | 0.29 (0.12-0.69) | 0.006 | 0.51 (0.32-0.81) | 0.005 |
| Male sex | 1.35 (0.89-2.05) | 0.16 | 1.80 (1.14-2.88) | 0.013 | 1.49 (0.65-3.43) | 0.35 | 0.91 (0.59-1.41) | 0.68 |
| Count of currently prescribed medications | 1.02 (0.97-1.08) | 0.44 | 0.96 (0.90-1.02) | 0.14 | 1.11 (1.02-1.22) | 0.020 | 1.00 (0.94-1.07) | 0.91 |
| Antihypertensive medications | 1.29 (0.82-2.03) | 0.27 | 1.27 (0.81-1.99) | 0.30 | 0.81 (0.35-1.94) | 0.63 | 0.95 (0.61-1.49) | 0.83 |
| Antidepressants | 0.87 (0.56-1.36) | 0.55 | 0.70 (0.42-1.15) | 0.16 | 0.50 (0.19-1.27) | 0.15 | 0.87 (0.53-1.41) | 0.56 |
| Antipsychotic medications | 2.03 (0.74-6.10) | 0.18 | 1.28 (0.42-4.09) | 0.67 | 1.46 (0.28-6.09) | 0.62 | 1.55 (0.48-4.72) | 0.45 |
| High anticholinergic burden | 0.75 (0.46-1.20) | 0.23 | 1.52 (0.90-2.55) | 0.11 | 1.22 (0.43-3.84) | 0.71 | 0.87 (0.54-1.42) | 0.58 |

\*Agree or strongly agree vs unsure, disagree or strongly disagree

Independent predictors of being willing to stop one or more of regular medicines were higher total involvement score (p=0.02), higher total burden score (p=0.001), lower total appropriateness score (p<0.001), lower total concerns about stopping score (p<0.001) and male sex (p=0.013) (Table 7). Independent predictors of total involvement score ≥4 (range 1-5) were lower total appropriateness score (p<0.001), higher total concerns about stopping score (p=0.009), age less than 65 years (p=0.006), and higher number of prescribed medications (p=0.02) (Table 7). Independent predictors of total appropriateness score ≥4 (range 1-5) were higher total involvement score (p=0.012), lower total burden score (p<<0.001), and age less than 65 years (p=0.005) (Table 7).

## Discussion

The paper presents the first assessment of patient attitudes towards deprescribing in the context of multimorbidity in the UK primary care setting. The study shows that even though most patients with multimorbidity are generally satisfied with their current medications, most (68% aged <65, 82% aged ≥65) would still be willing to stop one or more of their regular medicines if their doctor said it was possible. Polypharmacy was associated with a higher total burden score of using medications across age groups. Moreover, in those aged ≥65, patients with polypharmacy were more likely to be concerned with the appropriateness of their medications but, at the same time, more concerned about stopping them. This highlights the patient’s uncertainty about how medication changes may affect their health, even though ideas of stopping medications appear attractive. Unfortunately, the current clinical recommendations provide little guidance, adding pressure to both clinicians and patients.

Hypertension is the leading risk factor for cardiovascular and renal diseases and cognitive impairments with blood pressure-lowering medications widely used. However, while uncomplicated hypertension is typically asymptomatic, its treatments could cause adverse effects, including falls with uncertainty about optimal blood pressure targets in older people.(13) Indeed, our study shows that the use of antihypertensive medications, which were prescribed to 56% of those under 65 years and 76% of those ≥65 years, is of particular concern for patients. Their use was associated with a higher total burden score; patients whose medications included antihypertensive drugs were more willing to reduce the number of their medications and had a lower total appropriateness score.

Similarly, patients receiving antidepressants were less satisfied with their medications and had a lower total appropriateness score. There was an interesting observation that patients aged ≥65 whose medication included antidepressants were less likely to feel that their medications were appropriate and more concerned about stopping their medications. This is perhaps related to ongoing concerns about discontinuation effects that can occur on deprescribing antidepressants, although evidence from recent intervention trial shows this can be achieved successfully with support.(14) The latter was not observed about antihypertensive drugs, and could be due to the background presence of mental health issues (e.g., concerns of worsening mental health in case of medication discontinuation), reflecting challenges in making a decision and antihypertensive being perceived as a lower priority medication. This may reflect risk of relapse, but also confidence around their ability to manage without medication, lack of easy access to appropriate alternatives to medication (such as talking therapies), and their confidence around the competence of their general practitioner or pharmacist conducting a structured medication review to stop these safely, especially if these were initiated by a secondary care consultant psychiatrist or they have limited or no continuity of care within their GP practice (15). Qualitative studies show that fear of symptoms returning is a barrier to deprescribing in older patients.(16) Given the high prevalence of antidepressant use reported in populations with polypharmacy, understanding how best to engage these patients in discussions to achieve shared decision making to achieve deprescribing is needed in future studies.

Of interest, there was no significant difference in attitudes to deprescribing in people with and without frailty. This is important, as it sends the message the patients may not perceive the stigma of frailty as a factor justifying stopping their medications, which would be seen as equally important regardless of frailty status. This contrasts with the willingness to stop medication being highest among those aged ≥65 if receiving antihypertensive drugs, indicating that side effects affecting quality of life (which is typical of these drugs), rather than stigma of frailty are more important in attitudes for treatment choices. Living with multiple medical conditions is associated with poorer psychological well-being.(17) Such patients benefit from structured medication reviews to avoid overprescribing, particularly in the presence of cognitive impairment, when medication risks are more likely to outweigh the benefits.(8, 18) and reduced risk of hospitalization with falls or delirium, risk of functional decline and dementia and even death.(19, 20).

Our study demonstrates that patients who are satisfied with current medications are those who are involved in their prescribing. However, their satisfaction with treatments does not preclude their willingness to stop medications when appropriate. This should be considered during medication reviews.

*Limitations*: The study has several limitations. It is restricted to Evergreen Life PHR App users, which must be considered when generalizing the findings, including patients with lesser digital literacy and previous experience of systematic medications reviews. The antidepressant drug use was relatively high in the survey response, likely reflecting the multimorbid state of the study population, but might differ from populations in other regions. While the participation response rate is in keeping with typical response rates for other questionnaire surveys, there may be selection biases. For example, the participants included fewer patients receiving antipsychotic drugs than expected in a population with multimorbidity. When considering specific medication groups, the analysis cannot attribute the findings directly to specific drugs but rather to their presence among other drugs a patient uses. The reported views on deprescribing may differ from those when deprescribing is offered. In a small study of older patients’ or caregivers’ attitudes towards deprescribing in hospitals, a marked reluctance was observed towards trying deprescribing and a limited desire to be involved in shared decision-making.(21) This has been observed in deprescribing trials in other nations in the community, care home, or hospital setting for older patients.(22-25) Attempts to determine factors which may influence reluctance or refusal to participate in deprescribing potentially inappropriate medications have proven difficult to establish so far.(25)

## Conclusions

These findings suggest that patients taking multiple medications are generally satisfied with their medication but are willing to stop if recommended to do so, suggesting clinicians should consider proactively initiating de-prescribing discussions in patients with multimorbidity and polypharmacy. The presence of frailty does not increase likelihood of people wanting to stop medications; clinicians should proactively initiate deprescribing discussions and involve patients in the decision making process.

## Data Availability

All data produced in the present study are available upon reasonable request to the authors

## Acknowledgements

We are very grateful to all the participants that responded to the survey request and gave their time to this study.

## Acknowledgements

The Authors would like to thank Mary Braidley, Lee Campbell and Jack Higgins (formerly employees of Evergreen Ltd) for their initial contributions to this work, and to all the patients who kindly completed the rPATD survey. MG is part funded by the national institute of health and care research Applied Health Collaboration North West Coast. The views expressed here are those of the authors, not necessarily those of the National Institute for Health and Care Research nor the Department of Health and Care.

